# Neurocognitive Outcome after Pediatric Traumatic Brain Injury: Patient Subgroups with Diverging Outcome

**DOI:** 10.1101/2024.12.23.24319401

**Authors:** C.C. Kooper, M. Königs, M.E. Steenweg, M. Hunfeld, C.D. Scheurer, H.M. Schippers, W. Peper, A. Popma, J.B.M. van Woensel, D.R. Buis, H. Bruining, M. Engelen, J. Oosterlaan

**Affiliations:** Emma Neuroscience Group, Department of Pediatrics, Emma Children’s Hospital, Amsterdam UMC location University of Amsterdam, Amsterdam, The Netherlands; Amsterdam Reproduction and Development research institute, Amsterdam, The Netherlands; Emma Children’s Hospital Amsterdam UMC Follow Me program, Department of Pediatrics, Emma Children’s Hospital, Amsterdam UMC location University of Amsterdam, Amsterdam, The Netherlands; Department of Neurology, OLVG, Amsterdam, the Netherlands; Department of Pediatric Neurology, Erasmus MC, Sophia Children’s Hospital, Rotterdam, the Netherlands; Department of Pediatric Neurology, Spaarne Gasthuis Hospital, Haarlem and Hoofddorp, The Netherlands; Department of Neurology, St. Antonius Hospital, Utrecht and Nieuwegein, the Netherlands; Department of Pediatrics, St. Antonius Hospital, Utrecht and Nieuwegein, the Netherlands; Child and Youth Psychiatry, Emma Children’s Hospital, Amsterdam UMC location Vrije Universiteit Amsterdam & Levvel, Amsterdam, The Netherlands; Pediatric Intensive Care Unit, Emma Children’s Hospital, Amsterdam UMC location University of Amsterdam, Amsterdam, The Netherlands; Department of Neurosurgery, Amsterdam UMC location University of Amsterdam, Amsterdam, The Netherlands; N=You Centre, Emma Children’s Hospital, Amsterdam UMC location Vrije Universiteit Amsterdam & Levvel, Amsterdam, The Netherlands; Amsterdam Neuroscience research institute, Amsterdam, The Netherlands; Amsterdam Leukodystrophy Center, Department of Pediatric Neurology, Amsterdam UMC location University of Amsterdam, Amsterdam, The Netherlands

**Keywords:** Neurotrauma, Pediatrics, Clustering, Neurocognitive functioning

## Abstract

**Background and Objectives:** Traumatic brain injury (TBI) is the leading cause of acquired disability in children. Children with TBI are at risk of persistent deficits in neurocognitive functioning that affect daily life. However, neurocognitive outcomes are highly heterogenous and this heterogeneity is poorly understood. This study aims to investigate whether the heterogeneity in neurocognitive outcome can be reduced by distinguishing subgroups of children with distinct profiles of neurocognitive functioning, and to investigate whether these subgroups differ in demographic, premorbid and clinical characteristics.

**Methods:** This multicenter study included a consecutive cohort of children with mild to severe TBI and demographically matched neurologically healthy (NH) children. Seven neurocognitive domains were assessed six months post-TBI using computerized tests. The TBI and NH group were compared on the neurocognitive domains using t-tests. Results of the TBI group were subjected to cluster analysis to derive subgroups with distinct profiles of neurocognitive functioning. Resulting subgroups were compared on demographic, premorbid and clinical characteristics available at time of hospital visit.

**Results:** A total of 113 children with TBI and 113 NH children were included. The TBI group had lower performance than NH children on the neurocognitive domains Speed, Stability, Attention & Control, Verbal Working Memory and Visual Working Memory (.009≤*p*s≤.047, -.42≤*d*s≤-.29). Cluster analysis revealed four subgroups of patients with diverging neurocognitive outcome profiles. One subgroup was characterized by good outcome, whereas three subgroups had adverse outcome characterized by weak global outcome, weak visual-processing outcome or weak executive functioning outcome. The subgroups did not differ in clinical characteristics but did differ in demographic and premorbid characteristics. The weak global outcome subgroup had more premorbid behavioral problems, while the good outcome subgroup had higher socio-economic status than the other subgroups.

**Discussion:** This study indicates that children with mild to severe TBI exhibit neurocognitive deficits compared to matched controls at six months post TBI, among which subgroups of children with distinct neurocognitive outcome profiles exist. The neurocognitive outcome subgroups represent children with diverging severity and configuration of neurocognitive weaknesses. The subgroups with the diverging neurocognitive outcome profiles did not differ in clinical characteristics, highlighting the importance to consider other factors for the prognosis of neurocognitive outcome.

## Introduction

Pediatric traumatic brain injury (TBI) represents a significant public health concern, globally affecting over three million children per year.[1, 2] Children with TBI are at risk of neurocognitive deficits.[3, 4] These neurocognitive deficits often persist over time and can in turn affect other important domains of functioning, such as behavioral, emotional and academic functioning.[5–7] Consequently, neurocognitive functioning is regarded as a pivotal aspect of outcome after pediatric TBI.

Distinct differences in the nature and extent of neurocognitive consequences are observed among children with TBI, which are likely the result of the complex interplay between a range of biopsychosocial factors including premorbid functioning, the nature and severity of the injury, medical interventions and social environment of the child.[7–10] The limited understanding of the heterogeneous outcomes of TBI[11] in children challenges clinicians’ ability to recognize and manage children at risk of adverse development after TBI.[12]

Cluster analysis has shown promising potential to discern subgroups of TBI patients with distinct outcome. In adults, studies have identified patient subgroups with diverging outcome in symptom presentation,[13] behavioral functioning,[14] and neurocognitive functioning[15]. Similarly, clustering studies in children with TBI have identified subgroups with diverging outcome in behavioral,[16] neurocognitive[17–20] and academic functioning[21]. Moreover, subgroups of children with distinct IQ outcome also differed in behavioral functioning, with the subgroup most impaired in terms of intelligence also exhibiting the most behavior problems.[20] Consequently, the literature[22] indicates that cluster analysis can identify subgroups of children with diverging outcome. Distinguishing between these subgroups may aid in the understanding of factors that contribute to the existence of subgroups with distinct outcome which in turn may aid in outcome prognosis.

The few studies that used cluster analysis to understand the heterogeneity in neurocognitive outcome in pediatric TBI have important shortcomings. Existing studies have mainly focused on children with moderate-severe TBI,[16–19] while the vast majority of TBI cases concerns mild TBI[1] associated with abundant heterogeneity in outcome[23, 24]. Furthermore, existing studies exclusively used retrospective designs, lacking prospective registration of clinical parameters and involving patients with strong variation in time since injury, while recovery and post-injury development effects are well-known factors in neurocognitive outcome. Finally, existing studies have not investigated the relation between biopsychosocial factors and differences in neurocognitive outcome, which could improve our understanding of mechanisms that may lead to diverging neurocognitive outcome after TBI.

The current study aims to investigate the existence of subgroups of patients with diverging neurocognitive outcome in a sample of children with mild to severe TBI at six months post-injury using cluster analysis. In addition, the study addressed whether the discerned subgroups differed in demographic, premorbid and clinical factors available at time of their hospital visit. This study will contribute to our understanding of neurocognitive outcome heterogeneity and may provide insight into the factors that contribute to neurocognitive outcome.

## Methods

### Participants

This observational longitudinal multicenter study prospectively recruited children with TBI between December 2020 and December 2023 from two trauma-level 1 university medical centers and two general hospitals in the Netherlands (see[25] for study protocol). Inclusion criteria were: (i) diagnosis of TBI as determined by a pediatrician or pediatric neurologist; (ii) between 6 and 18 years old at time of outcome assessment; (iii) inhabitant of the Netherlands; (iv) fluent in Dutch; and (v) no diagnosis of a neurological disorder (other than TBI). Exclusion criteria were: (i) somatic disorders unrelated to TBI or severe psychomotor disability as this would interfere with assessment of (neurocognitive) functioning, and (ii) inability to understand test instructions. A total of 241 children were eligible for inclusion, of whom 20 were not reached and 98 declined participation. Reasons not to participate were as follows: not interested (*n*=26), no time (*n*=23), potential burden on child (*n*=24); while the remaining 25 eligible participants provided no reason for declining participation. Of all 123 children with TBI who provided consent for the study, 10 children did not participate in the outcome assessment (e.g. no time, personal circumstances) that took place six months post-injury.

The remaining 113 children with TBI were matched on a 1:1 ratio with 113 neurologically healthy (NH) children, recruited in a dedicated study designed to obtain neurocognitive and neurophysiological data in a representative sample of NH children. These children were recruited from various schools, social organizations, and sport clubs throughout the Netherlands between July 2021 and March 2024.[26] Inclusion and exclusion criteria for the NH group were identical to those of the TBI group, whereas children in the NH group were required to have no history of TBI as determined by parent report. Matching was based on age, sex and socio-economic status (SES) using the nearest neighbor method and logistic regression propensity scores (’MatchIt’[27] package).

#### Background information

Information about demographic characteristics (i.e., age, sex and SES) and medical history (i.e. clinical diagnoses) was collected using a digital parental questionnaire. SES was defined as the average level of parental education, ranging from no education (1) to postdoctoral education (8).[28] In terms of medical information, the questionnaire systematically assessed the presence of a broad range of disorders and comorbidities with specified examples for clarification. Medical history was then categorized into the presence of premorbid clinical diagnoses of learning disorders and psychiatric disorders.

#### Premorbid functioning

For children with TBI, information on premorbid functioning was collected using a digital parental questionnaire, administered as soon as possible after admission to the hospital (i.e., immediately after informed consent was acquired). Premorbid behavioral functioning was assessed using the ‘Strength and Difficulties Questionnaire’,[29] measuring emotional and behavioral problems in children (i.e. hyperactivity, emotional problems, problems with peers, behavioral problems, and prosocial behavior). The total problem score was used, where higher scores reflect more emotional and/or behavioral difficulties. Premorbid family functioning was assessed using the parent-rated ‘Questionnaire on Family Functioning for Parents’[30] which aims to assess the child’s family functioning (i.e. basic care, upbringing, social contacts, youth & experience, and partner relationship). The total score was used, where higher scores reflect fewer competency deficits in the parenting environment.

#### Injury-related characteristics

For children with TBI, information regarding injury mechanism and relevant clinical parameters during hospital stay were prospectively registered and extracted from the medical files in a structured manner (e.g. diagnosed abnormalities on computed tomography [CT], lowest Glasgow Coma Scale [GCS] score, duration of loss of consciousness [LOC], duration of post traumatic amnesia [PTA], admission to the pediatric intensive care unit and/or nursing ward).

### Neurocognitive outcome

Neurocognitive functioning was measured using the ‘Emma Toolbox for Neurocognitive Functioning’, an in-house developed composition of computerized tests based on well-established neuroscientific paradigms with proven validity and reliability.[26] Tasks used were: Attention Network Test,[31] Track & Trace Task,[32] Location Learning Test,[33] Rey Auditory Verbal Learning Test,[34] Klingberg Task,[35] and Digit Span[36]. See Appendix Table S1 for detailed descriptions of the neurocognitive tests used, and for an overview of the variables that were extracted from the data. Data obtained with these tasks were subjected to Principal Component Analyses to derive the neurocognitive domain scores Speed (i.e., processing speed, visuomotor precision and motor response inhibition), Stability (i.e., processing stability and processing consistency), Attention & Control (i.e., alerting attention, orienting attention, interference control speed and interference control precision), Memory (i.e., visual memory encoding, visual memory consolidation, verbal memory encoding and verbal memory consolidation), Verbal Working Memory (i.e., phonological loop and verbal central executive), Visual Working Memory (i.e., Visuo-spatial Sketchpad and Visual Central Executive) and Visuomotor Integration (i.e., Visuomotor Stability, Visuomotor Speed and Visuomotor Dynamic Integration). Higher neurocognitive domain scores reflect better performance. For all children, the scores were age-adjusted using regression weights obtained in the matched control group. The Emma Toolbox is programmed in Python using OpenSesame Software.[37]

### Procedure

The observational longitudinal multicenter study and the control study obtained approval from the Medical Ethical Committee Amsterdam University Medical Center (location AMC, reference number NL71283.018.19 and NL76915.018.21, respectively), and were registered at the International Clinical Trials Registry Platform (NL9051, NL9574, respectively). Written informed consent was provided by all children aged > 11 years old and parents of children aged < 16 years old. For outcome assessment, six months post-injury for the TBI group and after enrolment for the NH group, either a visit to the Emma Children’s Hospital or to our mobile laboratory (‘Emma Brain Bus’) was planned. All procedures were executed according to the Declaration of Helsinki.

### Statistical analysis

Statistical analysis was performed in R. The influence of outliers (values at >3 SD from the mean) was reduced by winsorizing (‘DescTools’[38] package) and missing data (<10%) was imputed (‘Mice’[39] package). To explore group comparability, the TBI and NH groups were compared on demographics and the prevalence of learning and psychiatric disorders as well as premorbid behavioral functioning, using t-tests and chi-square tests, where appropriate.

To investigate the impact of TBI on neurocognitive functioning, the TBI and NH group were compared using t-tests on the seven neurocognitive domain scores. Next, these domain scores were subjected to UMAP with k-means clustering[40] to determine subgroups of children with diverging neurocognitive outcome. UMAP has been shown to reduce the risk of using noise as a source for clustering.[41] The elbow method was used to determine the optimum number of clusters (i.e., subgroups of children). To aid in the interpretation of the resulting subgroups of patients with diverging outcome, each subgroup was compared to the NH group in terms of neurocognitive domain scores. To determine the risk of adverse neurocognitive outcome in each of the subgroups, we calculated the prevalence of children with a clinically relevant neurocognitive deficit (defined as performance <5^th^ percentile, corresponding to <1.65 z-score, on at least one neurocognitive domain) in each subgroup and compared this result to the prevalence in the NH group using chi-square tests. Similarly, each subgroup was also compared to the NH group on the average observed number of clinically relevant neurocognitive deficits using t-tests. To determine factors that may be related to neurocognitive subgroup membership, each subgroup was compared to all other subgroups together (e.g., 1 vs. 2,3,4) on demographic, premorbid and clinical factors using t-tests and chi-square tests, where appropriate. Demographic, premorbid and clinical factors with fewer than 10 occurrences were not used in these analysis.

Statistical analyses were two-sided with alpha set at .05 and multiple comparisons were accounted for by false discovery rate correction at the level of neurocognitive domain scores (*k*=7), demographics (*k*=3), premorbid functioning (*k*=2) and clinical characteristics (*k*=8). Cohen’s guidelines[42] for the interpretation of group differences were used, translating *d*=0.2 into small; *d=*0.5 into medium; and *d*≥0.8 into large effect sizes.

## Results

### Background information

As a result of the demographic matching procedure, children in the TBI and NH group did not differ in terms of sex, age at assessment and SES (see Table 1). The average level of SES reflected that both groups had on average parents with middle to higher education, which is representative for the Dutch population (i.e., 20% lower [1–4], 39% middle [5], 41% higher [6–8][28]). Regarding medical history and premorbid functioning, there were neither group differences in premorbid clinical diagnoses of learning or psychiatric disorders, nor in premorbid behavioral functioning (see Table 1).

**Table 1.** Study Sample Characteristics.

|  | TBI children<br>(n=113) | NH children<br>(n=113) | Statistic | *p* |
| --- | --- | --- | --- | --- |
| *Demographics* |  |  |  |  |
| Males, *n* (%) | 66 (58.4) | 51 (45.1) | $X^2 = 3.47$ | .06 |
| Age at assessment in y, *M* (SD) | 12.4 (3.7) | 11.7 (3.2) | $t = -1.73$ | .09 |
| Socio-economic status, *M* (SD) | 5.18 (1.13) | 5.37 (1.09) | $t = 1.32$ | .19 |
| *Premorbid diagnosed conditions and functioning* |  |  |  |  |
| Psychiatric disorder, *n* (%) | 6 (5.3) | 4 (3.5) | $X^2 = 0.38$ | .54 |
| Learning disorder, *n* (%) | 3 (2.7) | 7 (6.2) | $X^2 = 0.95$ | .33 |
| Behavioral functioning, *M* (SD) | 7.3 (4.7) | 6.6 (5.2) | $t = -0.98$ | .33 |
| *Injury-related characteristics* |  |  |  |  |
| Injury mechanism |  |  |  |  |
| Fall, *n* (%) | 50 (44.2) | - |  |  |
| Traffic accident, *n* (%) | 52 (46.0) | - |  |  |
| External object, *n* (%) | 11 (9.7) | - |  |  |
| Loss of consciousness (1-60 seconds), *n* (%) | 31 (27.4) | - |  |  |
| Loss of consciousness (>60 seconds), *n* (%) | 24 (21.2) | - |  |  |
| Post traumatic amnesia (1-60 minutes), *n* (%) | 33 (29.2) | - |  |  |
| Post traumatic amnesia (>60 minutes), *n* (%) | 20 (17.7) | - |  |  |
| *Initial assessment at ED* |  |  |  |  |
| Lowest Glasgow Coma Scale score | 13.3 (3.3) | - |  |  |
| Mild TBI (13-15), *n* (%) | 93 (82.3) | - |  |  |
| Moderate TBI (9-12), *n* (%) | 8 (7.1) | - |  |  |
| Severe TBI (3-8), *n* (%) | 12 (10.6) | - |  |  |
| Glasgow Coma Scale score < 15, *n* (%) | 45 (39.8) | - |  |  |
| Radiological findings |  |  |  |  |
| CT scan executed, *n* (%) | 82 (72.6) | - |  |  |
| CT abnormality, *n* (%) | 49 (43.4) | - |  |  |
| Skull fracture, *n* (%) | 33 (29.2) | - |  |  |
| Intracranial abnormality, *n* (%) | 40 (35.4) | - |  |  |
| Hemorrhage, *n* (%) | 34 (30.1) | - |  |  |
| Contusion, *n* (%) | 10 (8.8) | - |  |  |
| Mass effect, *n* (%) | 6 (5.3) | - |  |  |
| TAI, *n* (%) | 2 (1.8) | - |  |  |
| Neurosurgery, *n* (%) | 7 (6.2) | - |  |  |
| *Hospital admission* |  |  |  |  |
| Pediatric intensive care unit, *n* (%) | 24 (21.2) | - |  |  |
| Pediatric nursing ward, *n* (%) | 87 (77.0) | - |  |  |

*Note.* CT = computerized tomography; ED = emergency department; TAI = traumatic axonal injury; NH = neurologically healthy; TBI = traumatic brain injury.

### Injury-related characteristics

Children with TBI sustained the injury at the mean age of 11.4 years (*SD*=3.7, range: 5– 17), and included 66 boys (58.4%). Main trauma mechanisms were traffic accidents (*n*=52, 46.0%) and falls (*n*=50, 44.2%). Children had an average lowest GCS score of 13.3 (*SD*=3.3, range: 3–15), 55 children (48.7%) suffered from loss of consciousness at time of injury and 53 children (46.9%) had PTA as reported by the pediatrician or pediatric neurologist. Based on the lowest GCS score at time of the emergency department visit, 93 children (82.3%) had mild TBI, 8 children (7.1%) had moderate TBI and 12 children (10.6%) had severe TBI. Nonetheless, CT scans of 49 children (43.4%) revealed abnormalities of which 40 (35.4%) involved an intracranial abnormality (CT scan was conducted on 72.6% of the children). In addition, 87 children (77.0%) were admitted to the nursing ward, and 24 children (21.2%) were admitted to the intensive care unit. More details on injury-related characteristics are presented in Table 1.

### Neurocognitive functioning

#### Neurocognitive performance

Comparison of the TBI and NH groups on neurocognitive domain scores revealed small-sized group differences (Figure 1). The TBI group had poorer performance than the NH group on the neurocognitive domains Speed (*p*=.009; *d*=-.42), Stability (*p*=.009; *d*=-.38), Attention & Control (*p*=.009; *d*=-.39), Verbal Working Memory (*p*=.047; *d*=-.29) and Visual Working Memory (*p*=.009; *d*=-.39). No group differences were found in the domains Visuomotor Integration and Memory (*p*s≥.10, *d*s≤.23).

**Figure 1.**
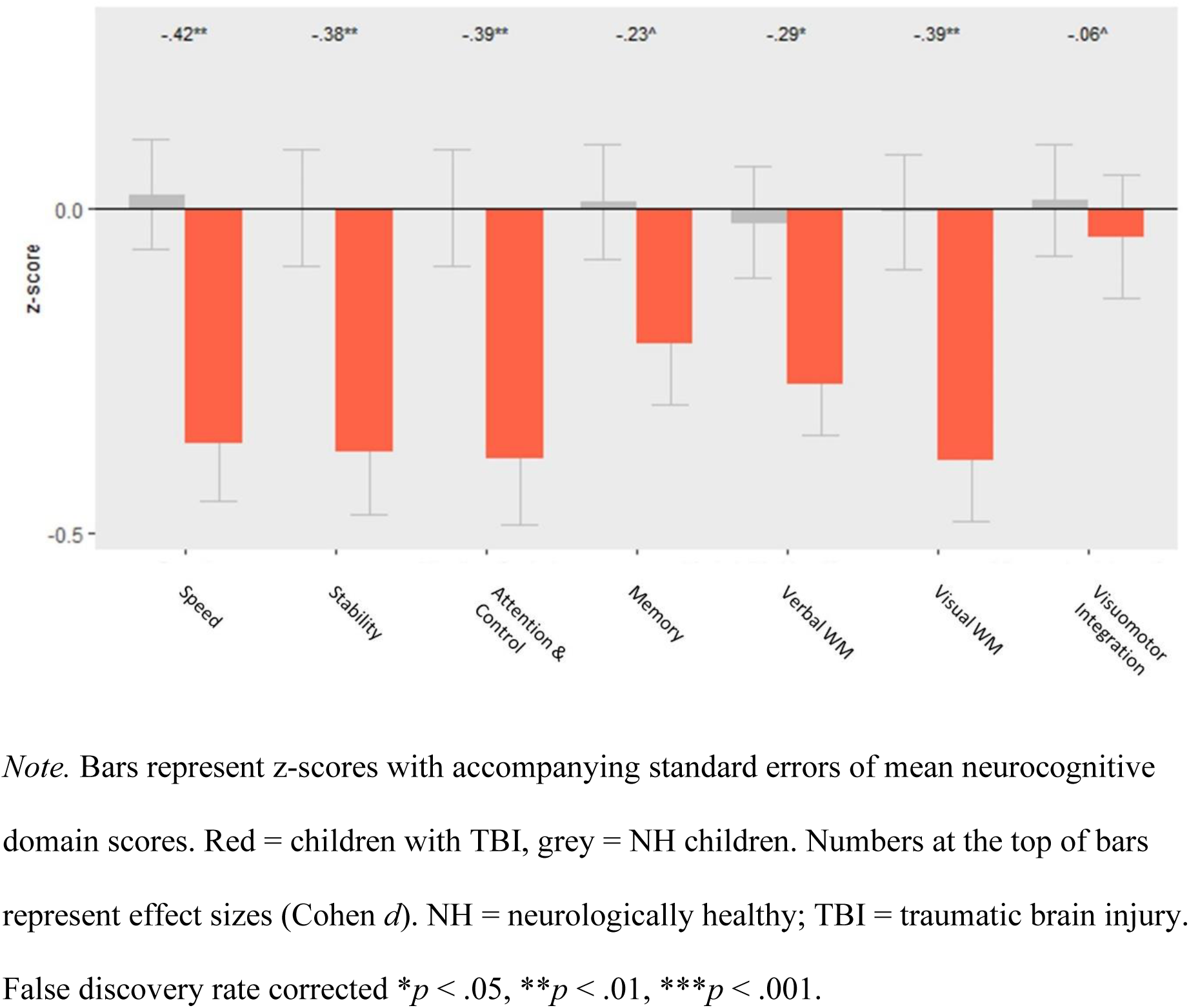
Neurocognitive Domain Scores of the TBI and NH group.

#### Neurocognitive subgroups

Cluster analysis showed that the optimal number of clusters (i.e., subgroups of children) to be extracted was four (Appendix Figure S1). Figure 2 displays the neurocognitive domain scores of the four derived subgroups with the NH group taken as reference group. Children in subgroup 1 (*n*=29, 25.7%) showed medium-sized differences compared to children in the NH group, with poorer performance on the neurocognitive domains Memory, Visuomotor Integration and Visual Working Memory (-.52≤*d*s≤-.65). Accordingly, this subgroup was labelled ‘weak visual-processing outcome’. Children in subgroup 2 (*n*=33, 29.2%) showed medium-sized and large-sized differences compared to children in the NH group, with poorer performance on the neurocognitive domains Speed, Stability, Attention & Control, Memory, Verbal Working Memory and Visual Working Memory (-.77≤*d*s≤-1.21). This subgroup was labelled ‘weak global outcome’. Children in subgroup 3 (*n*=24, 21.2%) showed medium-sized differences compared to children in the NH group, with better performance on the neurocognitive domains Memory and Verbal Working Memory (.57≤*d*s≤.71). This subgroup was labelled ‘good outcome’. Children in subgroup 4 (*n*=27, 23.9%) showed small-to medium-sized differences compared to children in the NH group, with poorer performance on the neurocognitive domains Speed, Stability, Attention & Control, Verbal Working Memory and Visual Working Memory (-.42≤*d*s≤-.67), whilst better performance on the domain Memory (*d=*.48). This subgroup was labelled ‘weak executive functioning outcome’.

**Figure 2.**
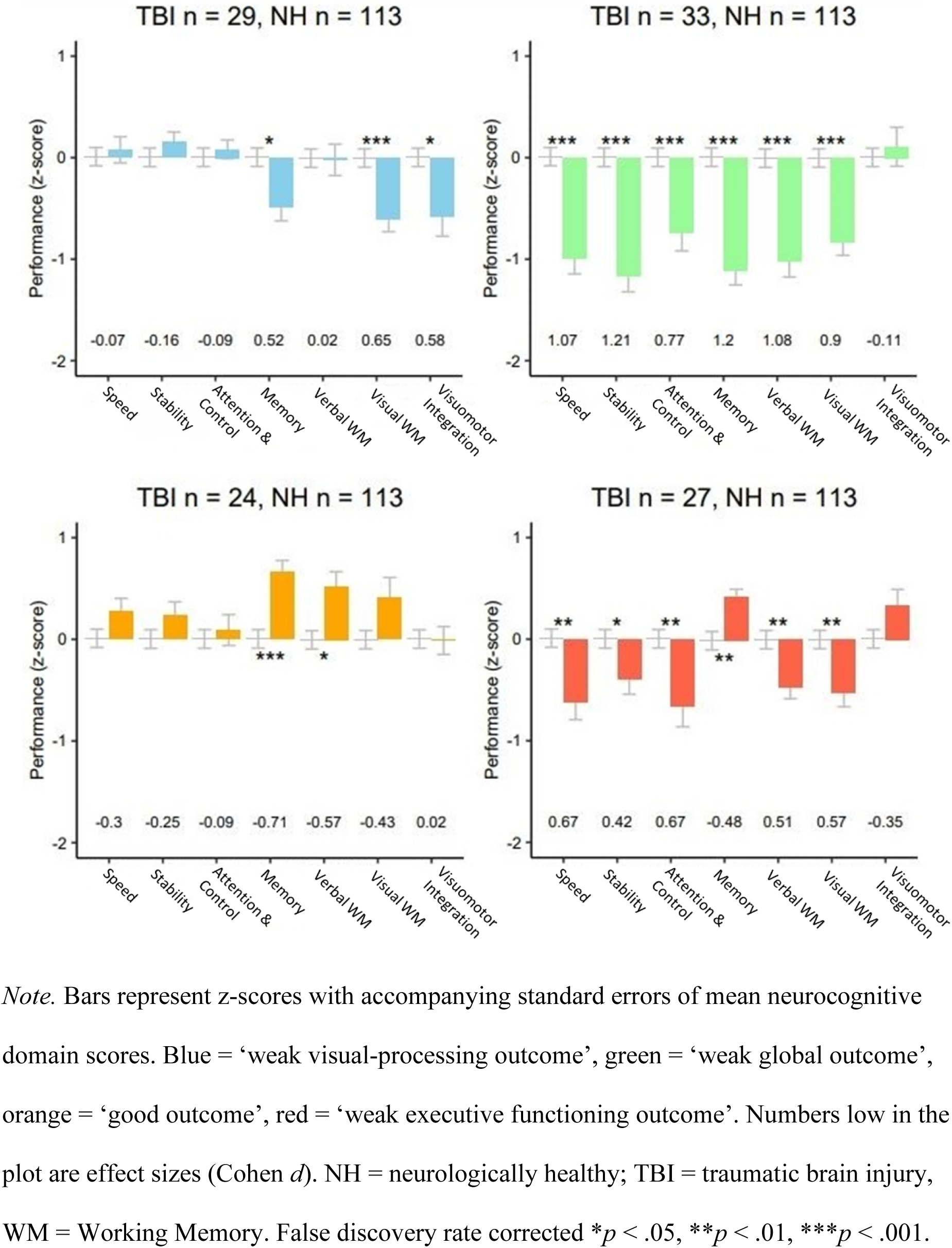
Neurocognitive Subgroups of Children with TBI with NH Group as Reference Group

The prevalence of children with a clinically relevant neurocognitive deficit (i.e., score <5^th^ percentile on at least one neurocognitive domain) is presented in Table 2 for each subgroup. Results showed children in the ‘weak global outcome’ subgroup had higher prevalence of at least one neurocognitive deficit (81.8%) than children in the NH group (29.2%), while the other subgroups did not differ from the NH group. Similarly, the average number of neurocognitive domains in which a deficit was present was higher among children in the ‘weak global outcome’ subgroup (*M*=1.52) compared to the NH group (*M*=0.37), but not in the other subgroups.

**Table 2.** Prevalence of Neurocognitive Deficits per Subgroup and for the NH Group.

|  | Subgroup 1<br>(n=29) | Subgroup 2<br>(n=33) | Subgroup 3<br>(n=24) | Subgroup 4<br>(n=27) | NH children<br>(n=113) | Contrast |
| --- | --- | --- | --- | --- | --- | --- |
| Neurocognitive deficit(s) in > 1 domain,<br>*n* (%) | 5 (17.2) | 27 (81.8) | 2 (8.3) | 11 (40.7) | 33 (29.2) | 2 > NH*** |
| Number of neurocognitive domain(s)<br>with deficit, *M* (SD) | 0.21 (0.49) | 1.52 (1.15) | 0.08 (0.28) | 0.48 (0.64) | 0.37 (0.64) | 2 > NH***<br>3 < NH** |

*Note.* Subgroup 1 = ‘weak visual-processing outcome’, Subgroup 2 = ‘weak global outcome’,
Subgroup 3 = ‘good outcome’, Subgroup 4 = ‘weak executive functioning outcome’. NH =
neurologically healthy. False discovery rate corrected $*p < .05$ , $**p < .01$ , $***p < .001$ contrasts with the NH group.

#### Characteristics at time of TBI per neurocognitive outcome profile

To determine factors that may be related to neurocognitive subgroup membership, each subgroup was compared to all other subgroups together on demographic characteristics, premorbid functioning, and injury-related characteristics (Table 3). Regarding demographics, children with TBI in the ‘good outcome’ subgroup on average had higher SES than children in the other subgroups. No significant differences in sex and age were found among the subgroups. With regard to premorbid functioning, children with TBI in the ‘weak global outcome’ subgroup on average exhibited more premorbid behavior problems than children in the other subgroups, while no significant differences were observed in premorbid family functioning. Regarding injury-related characteristics, no significant differences were found among the subgroups.

**Table 3.** Sample Characteristics Separately per Subgroup.

|  | Subgroup 1<br>(n=29) | Subgroup 2<br>(n=33) | Subgroup 3<br>(n=24) | Subgroup 4<br>(n=27) | Contrast |
| --- | --- | --- | --- | --- | --- |
| *Demographics* |  |  |  |  |  |
| Males, *n* (%) | 17 (58.6) | 22 (66.7) | 13 (54.2) | 14 (51.2) | NS |
| Age at assessment in *y*, *M* (*SD*) | 12.5 (3.1) | 12.2 (4.4) | 13.0 (2.8) | 12.2 (4.2) | NS |
| **Socio-economic status, *M* (*SD*)** | **5.2 (1.2)** | **4.8 (1.1)** | **5.8 (0.9)** | **5.0 (1.1)** | **3 > others\*\*\*** |
| *Premorbid functioning* |  |  |  |  |  |
| **Behavioral problems, *M* (*SD*)** | **6.2 (4.1)** | **9.1 (4.9)** | **6.9 (4.2)** | **6.6 (5.0)** | **2 > others\*** |
| Family functioning, *M* (*SD*) | 50.1 (10.3) | 49.7 (8.3) | 51.9 (11.6) | 51.8 (10.5) | NS |
| *Injury-related characteristics* |  |  |  |  |  |
| Loss of consciousness, *n* (%) | 11 (37.9) | 16 (48.5) | 13 (54.2) | 15 (55.6) | NS |
| Post traumatic amnesia, *n* (%) | 15 (51.7) | 14 (42.4) | 13 (54.2) | 11 (40.7) | NS |
| Lowest GCS score, *M* (*SD*) | 13.2 (3.7) | 12.4 (3.9) | 14.1 (1.8) | 13.7 (2.9) | NS |
| Mild TBI (GCS 13-15), *n* (%) | 25 (86.2) | 24 (72.7) | 21 (87.5) | 23 (85.2) | NS |
| Radiological findings on CT |  |  |  |  |  |
| Skull fracture, *n* (%) | 7 (24.1) | 12 (36.4) | 4 (16.7) | 10 (37.0) | NS |
| Intracranial abnormality, *n* (%) | 11 (37.9) | 14 (42.4) | 4 (16.7) | 11 (40.7) | NS |
| Hospital admission |  |  |  |  |  |
| Pediatric intensive care unit, *n* (%) | 5 (17.2) | 10 (30.3) | 1 (4.2) | 8 (29.6) | NS |
| Pediatric nursing ward, *n* (%) | 21 (72.4) | 25 (75.8) | 20 (83.3) | 21 (77.8) | NS |

*Note.* Subgroup 1 = ‘weak visual-processing outcome’, Subgroup 2 = ‘weak global outcome’, Subgroup 3 = ‘good outcome’, Subgroup 4 = ‘weak executive functioning outcome’. CT = computerized tomography; GCS = Glasgow Coma Scale; NS = not significant; TBI = traumatic brain injury. Text presented in bold reflect significant differences. False discovery rate corrected \* $p < .05$ , \*\* $p < .01$ , \*\*\* $p < .001$ .

## Discussion

This study investigated neurocognitive outcome using computerized tests after mild to severe pediatric TBI at six months post-injury. Children with TBI showed neurocognitive deficits compared to demographically matched peers, and cluster analysis revealed subgroups of children with distinct neurocognitive outcome profiles. These subgroups of children showed varying severity and configuration of neurocognitive weaknesses, and also varied in prevalence of clinically relevant neurocognitive deficits. Subgroup membership was related to demographic and premorbid characteristics, suggesting that these characteristics might predispose children to specific outcome profiles. In contrast, clinical features were not related to neurocognitive subgroup membership, indicating the limited value of clinical characteristics to predict the severity and nature of neurocognitive outcome. Neurocognitive outcome profiles may also aid future studies in identifying children at risk for adverse post-injury development and have the potential to guide effective follow-up care after emergency department treatment.

This study assessed neurocognitive functioning six months post-injury using an extensive and well-validated battery of computerized tests covering multiple neurocognitive domains. Comparisons between the group of children with (mild to severe) TBI and demographically matched NH peers showed that children with TBI had poorer performance in neurocognitive functions related to the speed and stability of information processing, attention control, and (visual and verbal) working memory. By utilizing a demographically matched NH control group with no differences in premorbid functioning as assessed by the prevalence of psychiatric and learning disorders or behavior problems, which were often not accounted for in previous studies, the observed group differences are likely to reflect the impact of TBI on neurocognitive functioning at six months post-injury. Neurocognitive functioning may still change over time due to late recovery effects or “growing into deficit” in more severe cases of TBI, highlighting the importance of follow-up studies to evaluate the trajectory of long-term neurocognitive outcome.[43] The findings of the current study are consistent with a large body of literature (for reviews see[3, 4]), and contribute by evaluating comprehensive neurocognitive profiles six months post-injury in a sample of children with mild to severe TBI as encountered at the emergency department. Inter-individual differences in neurocognitive outcome were further explored using cluster analysis to investigate whether the heterogeneity in neurocognitive outcome can be reduced by distinguishing distinct subgroups of children.

The results of the cluster analysis indeed revealed the existence of subgroups of children with distinct neurocognitive outcome. One subgroup, representing the minority of children with TBI (21%), exhibited good outcome. The remaining three subgroups had adverse outcome, together representing the majority (79%) of children with TBI. The subgroups that were characterized by adverse outcome, showed varying severity (medium-sized vs. large-sized effects) and diverging configurations of neurocognitive weaknesses. More specifically, one subgroup had global poor performance across neurocognitive domains, whereas the other subgroups had more specific weaknesses relating to either executive functioning or visual processing. Interestingly, the subgroups characterized by adverse outcome revealed weaknesses in neurocognitive domains that were not identified when examining the TBI group as a whole (i.e., in memory and visuomotor integration), indicating that certain neurocognitive weaknesses after TBI may go undetected in whole group comparisons. In addition, the results indicate that children in the subgroup with weak global outcome have a higher prevalence of clinically relevant neurocognitive deficits (i.e., performance <5^th^ percentile on at least one domain) compared to matched NH controls. Given previous research demonstrating a relation between neurocognitive functioning and long-term functional outcome (i.e., Extended Glasgow Outcome Scale) following TBI,[44, 45] these findings underscore the heightened risk for long-term adverse outcome, especially in this subgroup.

Our findings from the cluster analysis are in line with existing literature on adults with TBI, which has shown patient subgroups with differences in the severity and configuration of neurocognitive weaknesses relating to memory, speed, and executive functioning.[15] In pediatric TBI, this study enhances our understanding by providing a comprehensive evaluation across neurocognitive domains. The findings show that patterns in neurocognitive outcome are related to its favorability (i.e., good vs. adverse) and the configuration of adverse outcome (i.e., global weaknesses or specific weaknesses in functions relating to executive functioning or visual processing), rather than evaluating isolated domains (e.g., memory and attention,[17] or memory[18]). In addition, our findings indicate that differences in neurocognitive outcome profiles exist irrespective of varying recovery periods, providing a clearer understanding of outcome patterns at a specific post-injury timeframe (e.g., 6 months vs. 3-21 months[17]). Lastly, this study highlights neurocognitive outcome profiles in children with relatively milder injuries, providing insights that complement previous findings predominantly based on severe injuries[17,19].

We investigated the relation between subgroups of children with distinct neurocognitive profiles and demographic, premorbid and clinical factors. Children in the subgroup with weak global outcome also had higher levels of premorbid behavioral difficulties at time of the injury. This finding suggests that children with more premorbid behavioral difficulties are at greater risk of adverse neurocognitive outcome, in line with previous research in children with mild TBI.[46] Alternatively, children with premorbid behavioral difficulties may also have poorer premorbid neurocognitive functioning. The subgroup of children with good outcome also had higher SES (i.e., parental education), a demographic factor related to better neurocognitive functioning in healthy children.[47, 48] This finding suggests that SES may represent as a potential protective factor to adverse outcome, and complements a recent review[49] showing mixed findings between SES and neurocognitive outcome in TBI patients. Subgroups with specific neurocognitive weaknesses (i.e., weak executive functioning, weak visual processing) were not characterized by children with specific demographic, premorbid or clinical characteristics at time of the injury, indicating that these biopsychosocial factors do not seem to play a role in the prevalence of weaknesses in executive functioning or visual processing.

Interestingly, clinical features including TBI severity and diagnosed CT abnormalities were not significantly related to subgroup membership. It should be noted that more subtle relationships between clinical features and subgroup membership might have remained undetected, due the relatively small sample sizes of the neurocognitive outcome subgroups. Nevertheless, no strong relations were found, suggesting that clinical characteristics alone have limited value in predicting the severity and nature of neurocognitive outcome. This confirms previous findings using cluster analysis on variables from the acute stage, showing heterogeneous intellectual, adaptive and educational outcomes across the TBI severity spectrum and emphasizing the limited role of severity in determining outcome.[10] The lack of predictive factors for neurocognitive outcome underscore the need for routine follow-up with neurocognitive assessment to enable early detection of adverse neurocognitive outcome that may in turn affect post-injury development. Given the high prevalence of mild TBI, neurocognitive assessment of all children with TBI would not be feasible and highlights the need for robust predictive methods to identify those children at high risk of adverse neurocognitive outcome and requiring follow-up for early identification. Since previous pediatric studies have not extensively investigated the biopsychosocial factors contributing to diverging neurocognitive outcome profiles, further research is essential to understand how varying neurocognitive outcome affect a child’s development trajectory post TBI. Long-term monitoring is especially critical for children with outcome profiles that threaten future development (e.g., behavior, school performance, and employment prospects). Understanding the long-term outcomes of neurocognitive subgroups will help guide effective follow-up care after emergency department treatment.

This study has strengths and weaknesses. This observational multicenter study performed prospective registration of relevant clinical parameters for TBI as well as premorbid factors. In addition, we included children with a great variety in TBI injury severity, with the majority having mild TBI and representative of the TBI population seen in emergency departments.[24, 50] Another strength is the use of an extensive battery of well-validated neurocognitive computerized tests, that allowed to determine comprehensive neurocognitive outcome profiles, complemented by cluster analysis to better investigate the heterogeneity of neurocognitive outcome. However, it should be noted that statistical power to determine differences between subgroups was limited and the generalizability of findings to other cohorts of children with TBI remains unknown. Therefore, the findings of our study await replication in future research. Our findings also warrant further studies to investigate the nature and value of subgroups of children with distinct neurocognitive profiles. These studies should examine the relationship between neurocognitive subgroups and premorbid factors, characteristics related to injury and treatment, as well as whether diverging neurocognitive outcome also have differential repercussions for long-term development in behavioral and school functioning, adaptive functioning, quality of life and response to treatment.

## Conclusion

The results of this study show that children with TBI exhibit poorer neurocognitive outcome compared to matched peers, with subgroups of children with TBI showing distinct severity and configuration of neurocognitive weaknesses. Clinical characteristics show limited value to predict the severity and nature of neurocognitive outcome. The findings highlight patterns in the heterogeneity of neurocognitive outcome after TBI, and emphasize the need for further research to better understand mechanisms contributing to these differences in neurocognitive outcome and to identify children at risk for adverse post-injury development in behavioral, school, and adaptive functioning.

## Data Availability

All data produced in the present study are available upon reasonable request to the authors.

## Author Confirmation Statement

**Cece C. Kooper**: Conceptualization; Formal analysis; Investigation; Project administration; Writing - original draft; and Writing - review & editing. **Marsh Königs**: Conceptualization; Methodology; Software; Supervision; Funding acquisition; and Writing - review & editing. **Marjan E. Steenweg**: Resources; and Writing - review & editing. **Maayke Hunfeld**: Resources; and Writing - review & editing. **Nienke C.D. Scheurer**: Resources; and Writing - review & editing. **Herman M. Schippers**: Resources; and Writing - review & editing. **Arne Popma**: Writing - review & editing. **Job B.M. van Woensel**: Writing - review & editing. **Dennis R. Buis**: Writing - review & editing. **Hilgo Bruining**: Supervision; and Writing - review & editing. **Marc Engelen**: Conceptualization; Resources; Supervision; and Writing - review & editing. **Jaap Oosterlaan**: Funding acquisition; Supervision; and Writing - review & editing.

## Acknowledgements

We thank all participants and parents for their interest, time and contribution. Further, we thank all research assistants who helped with data collection.

