## Appendix Table S1 and Appendix Figure S1 for "Neurocognitive Outcome after Pediatric Traumatic Brain Injury: Patient Subgroups with Diverging Outcome"

**Table S1** Overview of the Neurocognitive Domains, Variables, Definitions and Tasks.

| **Domains** & Variables | Description | Definition | Task | Descriptives |
|---|---|---|---|---|
| **Speed (domain score*)** | | | | **M = 0**<br>**SD = 0.9**<br>**Range: -2.8 to1.2** |
| Processing Speed | The speed of responding to target appearance | Mean reaction time (ms) on trials with neutral targets. | Attention Network Test | M = 587.8 ms<br>SD = 141.8<br>Range: 354 to 1055 |
| Visuomotor Precision | The precision of proactive visuomotor tracking | The mean distance (in pixels) between the target and the mouse cursor in the structured condition across speed levels. | Track & Trace task | M = 93.6 pixels<br>SD = 68.5<br>Range: 29 to 355 |
| Motor Response Inhibition | The speed of disengaging from an ongoing motor response and engaging an adapted motor response | The mean time (ms) after a path change of the stimulus in the unstructured condition at which the peak distance is observed between the target and the mouse cursor. | Track & Trace task | M = 322.9 ms<br>SD = 79.3<br>Range: 229 to 649 |
| **Stability (domain score)** | | | | **M = 0**<br>**SD = 1.0**<br>**Range : -2.9 to 1.6** |
| Processing Stability | The variability of responding to target appearance | Standard deviation of *Processing Speed*, expressed as a percentage of *Processing Speed*. | Attention Network Test | M = 18.4%<br>SD = 4.8<br>Range: 8 to 32 |
| Processing Consistency | The consistency of responding to target appearance | The mean of the exponential curve in Ex-Gaussian analysis (Massidda, 2013) of the *Processing Speed* distribution including all trials, corrected for trial type, expressed as a percentage of the *Processing Speed*. | Attention Network Test | M = 16.6%<br>SD = 6.1<br>Range: 5 to 35 |
| **Attention Control (domain score)** | | | | **M = 0**<br>**SD = 1.0**<br>**Range: -3.4 to 1.6** |
| Alerting Attention | The gain in processing speed by temporally activating attention | The difference in mean reaction time (ms) between trials with a central cue and trials without a cue, expressed as a percentage of the latter. | Attention Network Test | M = -3.5%<br>SD = 5.4<br>Range: -12 to 18 |
| Orienting Attention | The gain in processing speed by temporarily activating spatially orienting attention | The difference in mean reaction time (ms) between trials with spatial and central cues, expressed as a percentage of the latter. | Attention Network Test | M = -8.3%<br>SD = 5.1<br>Range: -20 to 10 |
| Interference Control Speed | The speed of suppressing irrelevant information | The difference in mean reaction time (ms) between trials with incongruent and congruent targets, expressed as a percentage of the latter. | Attention Network Test | M = 23.3%<br>SD = 8.3<br>Range: -7 to 58 |
| Interference Control Precision | The accuracy of suppressing irrelevant information | The difference in accuracy (number of errors) between trials with incongruent and congruent targets, expressed as a percentage of the latter. | Attention Network Test | M = -8.4%<br>SD = 8.5<br>Range: -38 to 12 |
| **Memory (domain score)** | | | | **M = 0**<br>**SD = 0.9**<br>**Range -3.2 to 1.4** |

| | | | | |
|---|---|---|---|---|
| Visual Memory Encoding | The ability to encode visual information in short-term memory | The sum of correct displacements over five direct recall trials. | Location Learning Test | M = 35.6<br>SD = 23.5<br>Range: 0 to 119 |
| Visual Memory Consolidation | The ability to consolidate visual information in long-term memory | The number of correctly recognized positions expressed as a percentage of the number of unique items presented in the direct recall trial. | Location Learning Test | M = 87.1%<br>SD = 15.4<br>Range: 30 to 100 |
| Verbal Memory Encoding | The ability to encode verbal information in short-term memory | The sum of correct words recalled over the five direct recall trials. | Rey Auditory Verbal Learning Test | M = 45.8<br>SD = 11.1<br>Range: 16 to 69 |
| Verbal Memory Consolidation | The ability to consolidate verbal information in long-term memory | The number of correctly recognized words expressed as a percentage of the number of unique words presented in the direct recall trial. | Rey Auditory Verbal Learning Test | M = 97.7%<br>SD = 6.7<br>Range: 46.7 to 100 |
| **Visual Working Memory (domain score)** | | | | **M = 0**<br>**SD = 1.0**<br>**Range: -1.9 to 3.8** |
| Visuo-spatial Sketchpad | The capacity of encoding visual information in short-term memory. | Performance in the forward condition, as determined by the number of correct responses multiplied by the span of the item with the last correct response. | Klingberg Task | M = 76.6<br>SD = 36.6<br>Range 10 to 187 |
| Visual Central Executive | The capacity of the central executive to manipulate visual information in short-term memory. | The change in performance between the backward and the forward condition, expressed as a percentage of the latter. | Klingberg Task | M = -5.0%<br>SD = 99.5<br>Range: -92 to 800 |
| **Verbal Working Memory (domain score)** | | | | **M = 0**<br>**SD = 0.9**<br>**Range -1.9 to 2.1** |
| Phonological loop | The capacity of encoding verbal information in short-term memory. | Performance in the forward condition, as determined by the number of correct responses multiplied by the span of the item with the last correct response. | Digit Span | M = 43.6<br>SD = 20.2<br>Range: 2 to 104 |
| Verbal Central Executive | The capacity of the central executive to manipulate verbal information in short-term memory. | The change in performance between the backward and the forward condition, expressed as a percentage of the latter. | Digit Span | M = -32.0%<br>SD = 43.5<br>Range: -200 to 90 |
| **Visuomotor Integration (domain score)** | | | | **M = 0**<br>**SD = 0.9**<br>**Range: -2.7 to 2.0** |
| Visuomotor Stability | The variability of proactive visuomotor tracking. | The standard deviation of the mean distance (in pixels) between the target and the mouse cursor in the structured condition, expressed as a percentage of the mean distance (across speed levels). | Track & Trace Task | M = 78.0%<br>SD = 12.8<br>Range: 58 to 125 |
| Visuomotor Speed | The resistance to increasing speed requirement in visuomotor tracking. | The gain in mean distance (in pixels) between the fastest condition and the slowest condition, expressed as a percentage of the latter (across structured and unstructured conditions). | Track & Trace Task | M = 219.9%<br>SD = 65.5<br>Range: 78 to 373 |

| | | | | |
|---|---|---|---|---|
| Visuomotor Dynamic Integration | The precision of reactive visuomotor tracking. | The gain in mean distance (in pixels) between the unstructured condition and the structured condition, expressed as a percentage of the latter (across speed levels). | Track & Trace Task | M = 58.8%<br>SD = 46.2<br>Range: 11 to 439 |

*Note*. Experimental procedures (‘Tasks’) that have been applied to generate test scores targeting specific neurocognitive functions (‘Variables’), which in turn were clustered using principal component analysis to retrieve overarching scores representing neurocognitive domains (‘Domains’). *Domain scores represent z-scores of neurologically healthy children. ms = milliseconds.

**Figure S1.** Elbow Method to Decide on Optimal Number of Clusters (i.e., Subgroups).

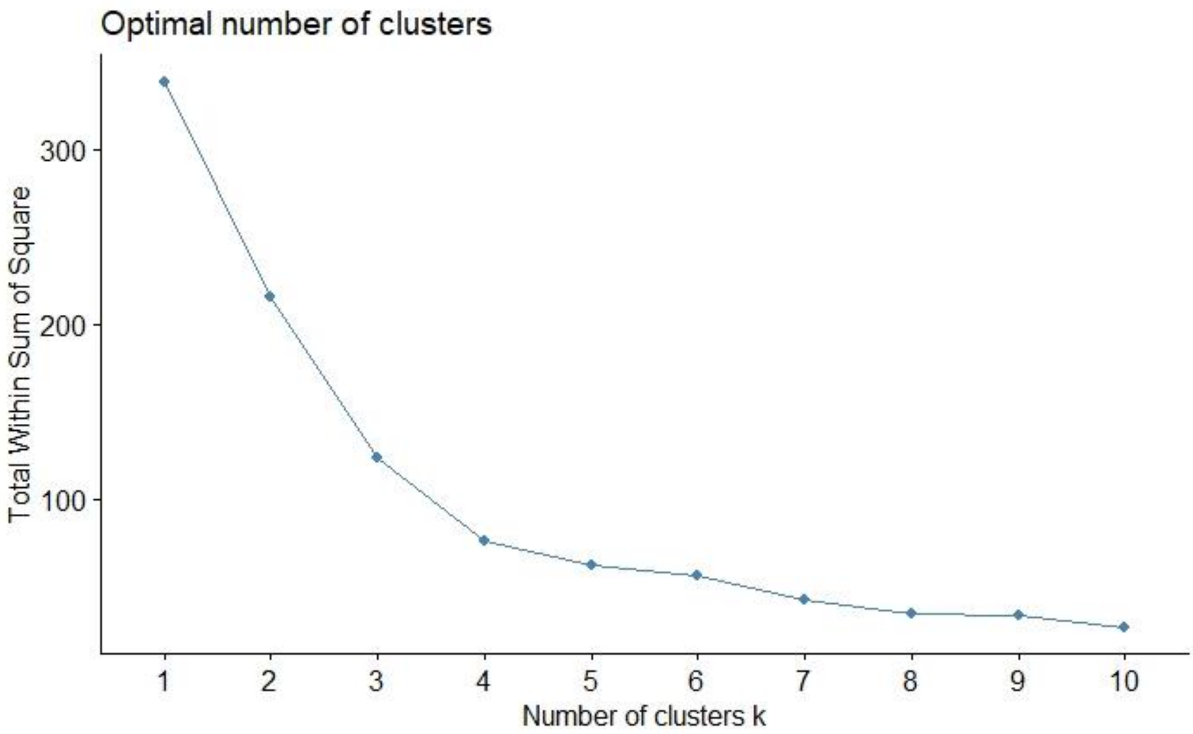
